# MTPpilot: an interactive software for visualization of NGS results in molecular tumor boards

**DOI:** 10.1101/2022.02.09.22270751

**Authors:** A. Kahraman, F. M. Arnold, J. Hanimann, M. Nowak, C. Pauli, C. Britschgi, H. Moch, M. Zoche

**Affiliations:** Department of Pathology and Molecular Pathology, University Hospital Zurich (USZ); Swiss Institute of Bioinformatics (SIB); Faculty of Medicine, University of Zurich (UZH); Department of Medical Oncology and Hematology, Comprehensive Cancer Center Zurich, University Hospital Zurich (USZ)

## Abstract

Comprehensive targeted Next Generation Sequencing (NGS) panels are routinely used in modern molecular cancer diagnostics. In molecular tumor boards the detected genomic alterations are often discussed to decide the next treatment options for the cancer patient. With the increasing size and complexity of NGS panels, the discussion of these results becomes increasingly complex, especially if they are reported in a text-based form, as it is the standard in current molecular pathology. We developed the Molecular Tumor Profiling pilot (*MTPpilot*) software to enable an efficient and quick analysis and visualization of complex NGS results, thanks to a combination of automated annotations and interactive tools. The software is tailored for the use at molecular tumor boards to aid clinical decision making. It is freely available as a web-application at https://www.mtppilot.org.

## INTRODUCTION

Next Generation Sequencing (NGS) has become a key cornerstone for today’s clinical decision-making in oncology. Comprehensive NGS cancer panels are widely used and constitute an integral part of modern molecular diagnostics [1]. Results of NGS cancer panels are often discussed at molecular tumor boards, where oncologists, molecular pathologists and staff scientists decide on the best treatment options for a patient [2-6]. With the transition from small panels covering hotspot regions of actionable genes to larger comprehensive panels, NGS results have become more challenging to understand and interpret [7]. Typically, only a subset of the detected alterations are well-known actionable mutations. Most alterations are scarcely described or are of unknown significance [8]. Different institutions and private companies have tackled this issue by providing automated molecular reports that classify alterations according to their pathogenicity and evidence in the literature or cancer databases. While these reports greatly help the physicians to interpret NGS results, they still feature several drawbacks. They generally lack visualization of the results and are very text-focused, making them often very cumbersome to read, especially in the case of many alterations. In addition, text based reports are less convenient for a dynamic discussion at molecular tumor boards, where interactive analysis of the results is preferred.

Several solutions tackling this problem have been recently developed, such as cBioPortal, Swiss-PO or AML Varan. cBioPortal is aimed to visualize NGS results of large research cohorts like The Cancer Genome Atlas (TCGA) [9, 10]. Swiss-PO is a tool for structural analysis of mutations in common cancer genes [11]. AML Varan is a very comprehensive solution, which offers the complete workflow from raw data analysis to medical report generation, but lacks the interactive visualization of results for molecular tumor boards [12]. Local software solutions have also been developed by commercial providers, but require access fees. With the increasing complexity of NGS data, a simple comprehensive software solution is required that integrates all aspects of NGS data analysis in the context of precision oncology. The solution should allow users (i) to easily upload their data, (ii) to classify alterations according to publicly available databases, (iii) to visualize alterations with interactive tools and (iv) provide all these features in a fully automated manner and in real-time, to allow an efficient discussion at molecular tumor boards.

To address these points, we developed the Molecular Tumor Profiling Pilot (*MTPpilot*) software in close collaboration with oncologists and pathologists within the molecular tumor board of the University Hospital Zurich. *MTPpilot* is tailored for various comprehensive panels and provides a set of fully automated annotations and tools for the interpretation of genomic alterations from several different perspectives. To the best of our knowledge, this is the first freely available software that offers a holistic set of tools to easily analyze NGS data and to aid clinical decision making at molecular tumor boards. The *MTPpilot* is freely available as a web service.

## RESULTS

### Data upload and graphical interface

Users can upload files either in tab separated value (tsv) format, or enter the mutational data manually via a table upload form. The tsv format is specified by an example file available in the csv upload section, and the upload is limited to 200 lines of alterations. After uploading, the user is able to visually inspect the data in a table view, and errors are highlighted to the user. This visual feedback was designed for a better user experience, especially in the context of large datasets. Conventions for how to annotate nucleotide and amino acids changes are given in a tutorial section. After the successful submission of the data, the user is redirected to the graphical interface of the *MTPpilot* software. An overview of the graphical interface and its features is given in Figure 1.

**Figure 1:**
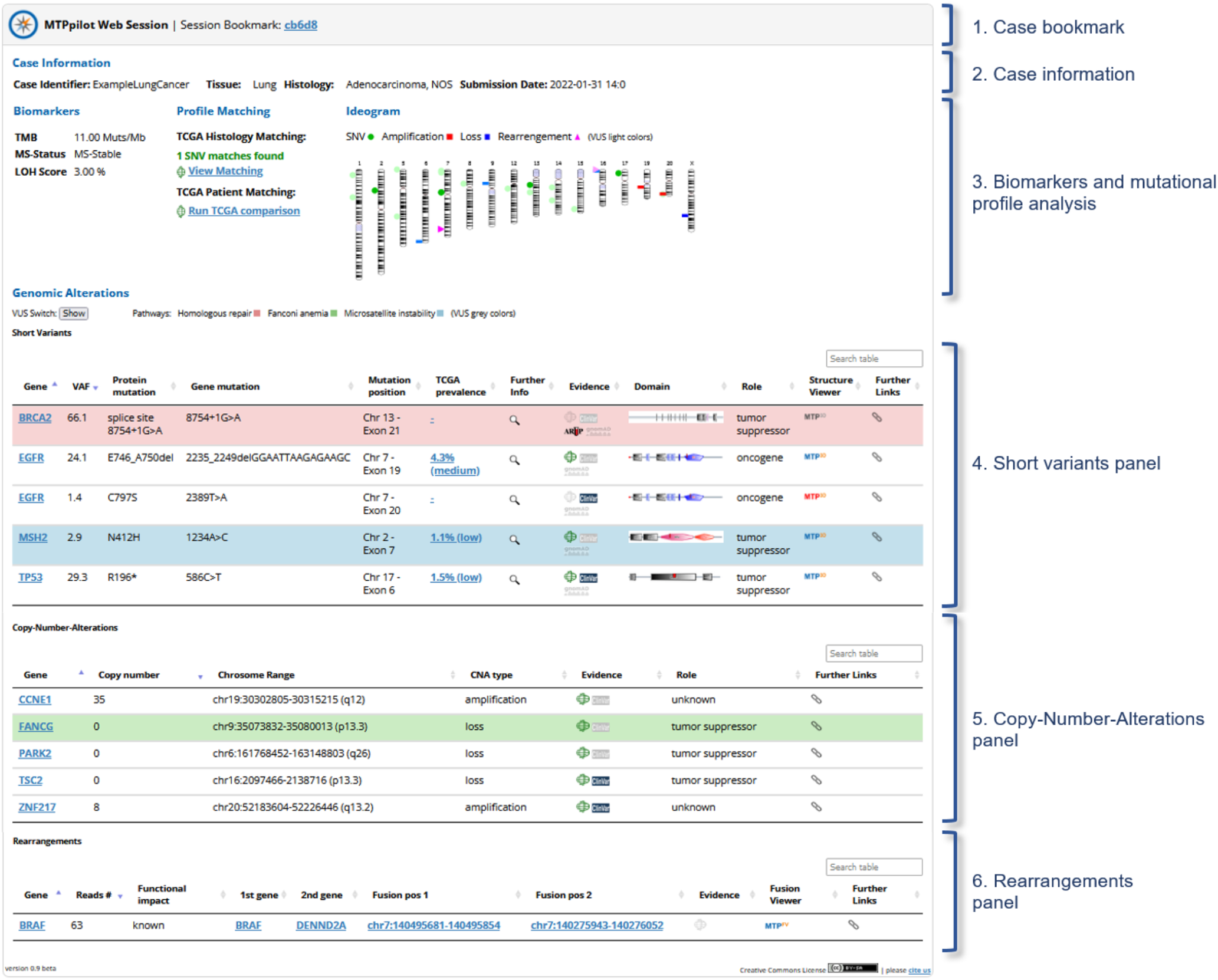
The *MTPpilot* graphical interface. The interface is subdivided into: (i) a case specific bookmark; (ii) a header with case information; (iii) a mutational profile analysis panel, which features the biomarkers provided by the user, profile and patient matching tools, and a plot of the mutations on the affected chromosomes (ideogram); (iv) a short variant panel, offering several tools to analyze short nucleotide variants; (v) a copy-number-alterations panel; (vi) and a panel for the inspection of rearrangements, such as fusions or truncations.

### Case Bookmark and Case Information

The case bookmark can be used by the user to retrieve the current session after he leaves the *MTPpilot* website, without having to re-upload the data. The bookmark is valid for 30 days, after which the data will be automatically deleted. The case information displays the case identifier, the tissue and the histology provided by the user, as well as the submission time and date.

### Genomic Biomarkers and Mutational Profiles

The genomic biomarkers and mutational profile panel (Figure 2) displays the tumor mutational burden (TMB) [13], Microsatellite (MS) status [14] and loss of heterozygosity (LOH) score [15]. The profile matching section offers two matching tools: (i) a TCGA histology matcher and (ii) a TCGA patient matcher. In the TCGA histology matcher, short nucleotide variants (SNVs) are matched against the top 20 mutated genes for a preselected cancer histology in the TCGA dataset [10]. Via a pop-up, a histogram highlights the match between TCGA and user provided altered genes (Figure 2A). This allows the user to assess how well the provided mutations fit to the selected histology. In the TCGA patient matcher tool, all alterations are matched against the mutations of each TCGA sample. Matched samples are listed in table form ordered by similarity, and the tissue frequencies of matched samples are reported graphically (Figure 2B). This allows the user to identify TCGA samples with similar alteration profiles. These tools are especially beneficial for checking and predicting cancer histologies, e.g. in the case of cancers of unknown primary (CUPs). All mutations are plotted on an ideogram showing the alterations by type (single nucleotide variants, amplifications, losses and rearrangements) on the affected chromosomes (Figure 1). The ideogram allows grasping the mutational landscape of a case, identifying for example highly mutated tumors or homologous recombination deficiency (HRD) in the case of many copy number alterations.

**Figure 2:**
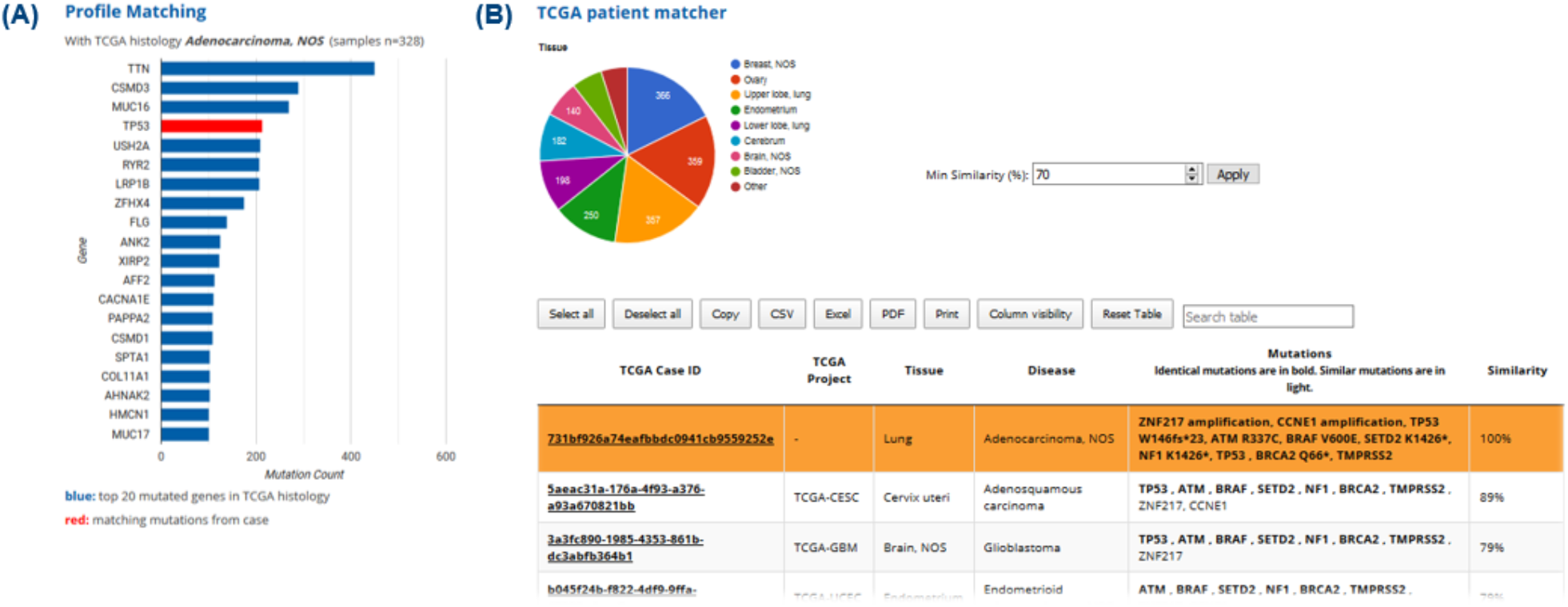
Profile analysis with TCGA data. (A) In the profile matching tool the altered genes are matched against the top 20 most frequently altered genes of the TCGA histology provided by the user. Matched genes are highlighted in red. (B) Alterations are matched against each TCGA sample in the TCGA patient matcher tool. The most similar samples are displayed in table form, and the tissue distribution of these samples are reported in a pie chart. The user can manually set the minimum similarity threshold for matching.

### Short Variants Analysis

The short variants analysis (SNV) panel offers comprehensive annotations and several tools for inspection of SNV, insertions, deletions (indels) and frameshifts (Figure 3A). Variants of unknown significance (VUS) can be displayed or hidden by using the toggle switch at the top of the panel. This is useful to avoid overcrowding the panel, in particular for cases with high mutational burden. The first five columns provide standard annotations such as protein sequence, coding sequence and exon annotations with outlinks to bioinformatic databases (Figure 3A column 1 to 5). The TCGA prevalence tool shows a histogram of the most frequently mutated amino acid positions within a gene in the TCGA dataset (Figure 3C and Figure 3A column 7). The tool helps to determine, whether a user provided alteration lies in a mutational ‘hot-spot’. The evidence column provides information if an alteration is described in the TCGA, ClinVar [16], ARUP [17] or gnomAD [18] databases. If no evidence is available, the database link is greyed out (Figure 3A column 8). The SMART [19] domain viewer shows the position of the alteration within the secondary structure of the protein and conserved domains (Figure 3D and Figure 3A column 9). The *MTP3D* tool displays the special coordinates of the altered amino acid on protein structures (Figure 3B and Figure 3A column 11). A red icon indicates the availability of a Protein Data Bank (PDB) [20] structure from the with the mutated amino acid, a blue icon indicates a PDB structure covering the mutated amino acid position, and a grey icon indicates a PDB structure lacking the mutated amino acid position. If no PDB structure is available, no icon is shown. The user can choose different structures from the PDB database, and can choose between different visualization forms.

**Figure 3:**
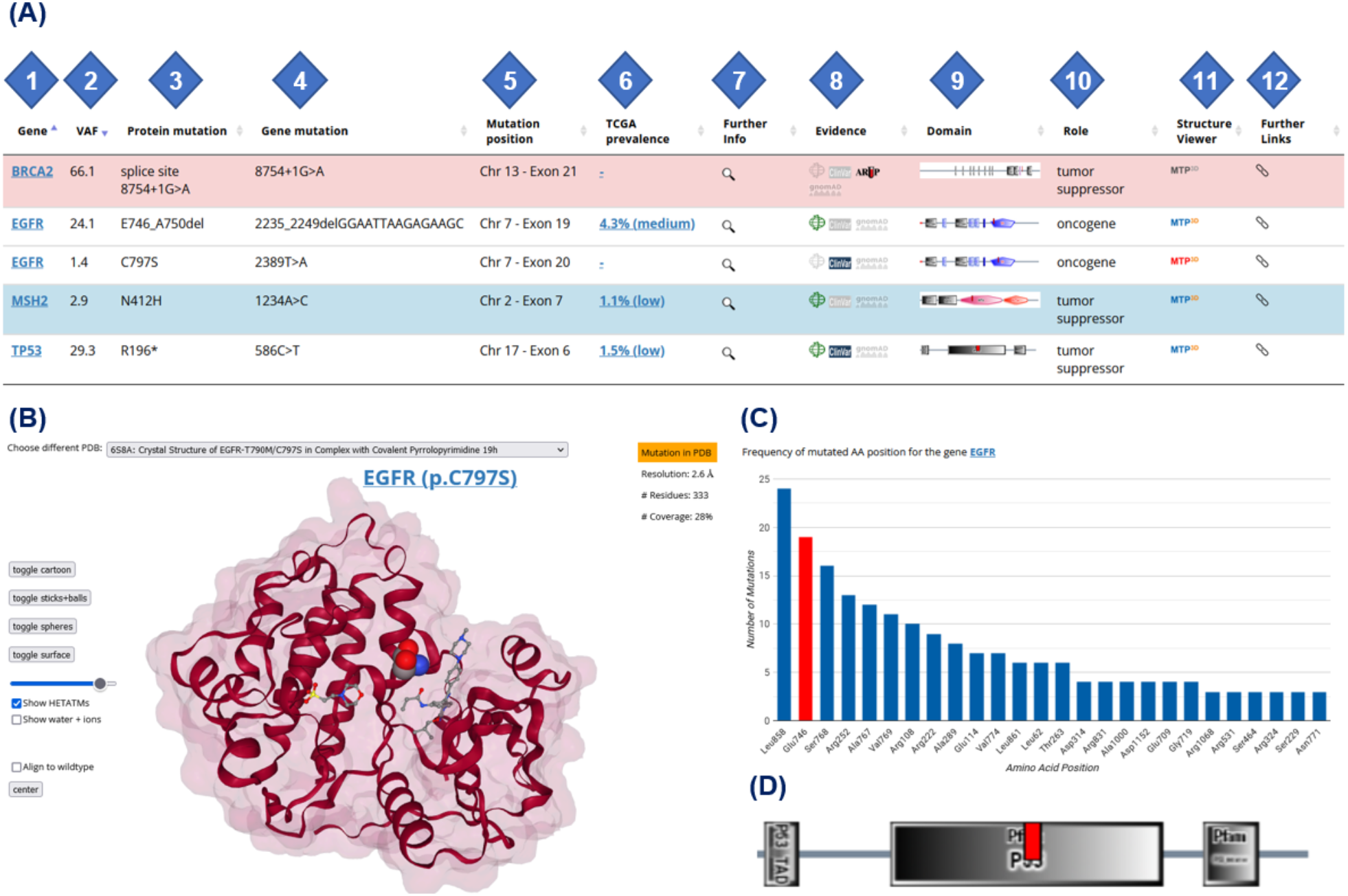
Features of the single nucleotide variant panel. (A) The different columns of the panel comprising annotation and links to interactive tools are highlighted by numbers: (1) protein name link to the UniProt entry; (2) variant allele frequency provided by the user; (3) protein mutation; (4) nucleotide alteration; (5) chromosome and exon number of the alteration; (6) TCGA amino acid position prevalence tool; (7) further informations such as chromosomal coordinates or amino acid conservation score; (8) evidence inspection tool comprising the databases TCGA, ClinVar, gnomAD and for BRCA genes ARUP; (9) visualization of the alteration (red tag) within the 2-dimensional SMART domains of the protein; (10) biological role of the gene according to KEGG pathways; (11) link to *MTP3D* viewer tool for analysis of alterations on three-dimensional protein structures; (12) further links to other databases and NGS panels. (B) The *MTP3D* viewer tool displays the altered amino acid as a sphere model on the structure with the highest resolution of the affected protein in cartoon rapresentation. Different views such as stick & balls, spheres or surfaces can be chosen, as well as other structures from the PDB database. In this example, the EGFR alteration C797S is displayed on PDB ID 6SBA (C) The TCGA prevalence tool shows the frequency of alterations at a given amino acid position for the affected protein, and highlights the alteration provided by the user in red. In this example, the EGFR E746 position was altered and is highlighted as one of the 20 most common mutated amino acid positions of P53. (D) The SMART domain tool displays the location of the short nucleotide variant (red line) within the protein domains. In this example the TP53 R196* terminating mutation is shown within the protein tyrosine kinase domain of BRAF.

The combined information from all tools of the SNV panel enables an integrated analysis, and a more confident decision-making in characterizing an alteration, in particular in case of less known alterations or genes.

### Copy-Number Variants Panel

The copy number analysis panel (Figure 1) displays information such as the chromosomal location, as well as the evidence in the TCGA dataset and in ClinVar. This section is useful in conjunction with the ideogram, where the copy-number variants panel shows the copy number changes while the ideogram highlights the distribution of gains and losses over the affected chromosomes.

### Rearrangement Analysis Panel

The rearrangement analysis panel (Figure 1) displays the chromosomal coordinates of the fusion breakpoints, the TCGA evidence viewer and a link for the *MTPfusion* viewer tool. The TCGA evidence viewer shows the fusion partners for the rearranged gene in the TCGA dataset, as well as the tissue frequencies, in which the rearranged gene was observed. The *MTPfusion* viewer gives a graphical representation of the resulting fusion event. The user can choose between different protein transcripts and sequence orientations (Figure 4). This automatic annotation avoids laborious manual annotation of different rearrangement possibilities, and is especially helpful in the context of gene fusions and truncations.

**Figure 4:**
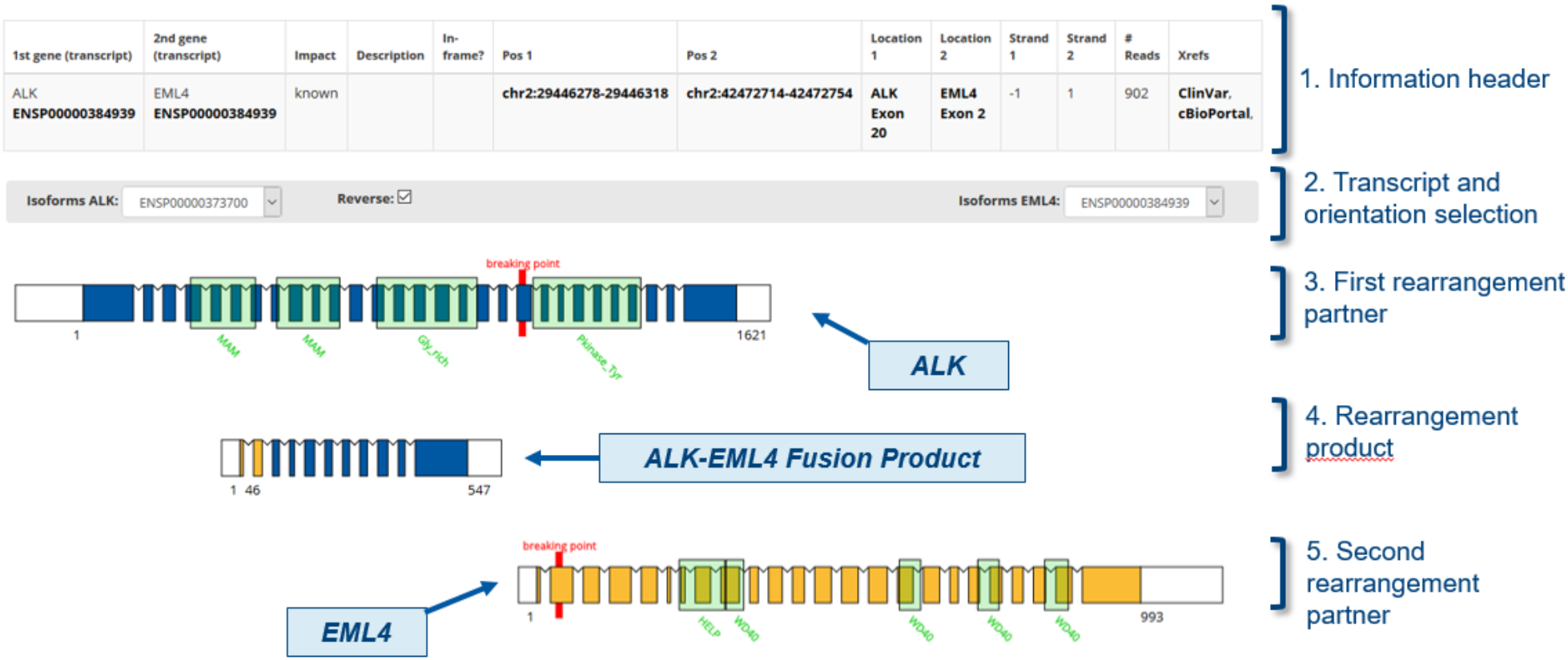
The *MTPfusion* viewer tool. The tool displays a case of a canonical *ALK-EML4* Fusion. Breakpoints of the involved genes provided by the user are highlighted in red. The tool allows to change the isoform of the two genes involved in the rearrangement. The orientation of the rearrangement can be flipped with the reverse selector.

## CONCLUSIONS

NGS data has become bigger and more complex, making the analysis of variants more challenging. Often a consultation of different databases and a combination of bioinformatic tools are used to characterize cancer alterations and to assess their pathogenic impact. However, no software is so far available that integrates all necessary information in one solution. The *MTPpilot* fills this gap by automatically annotating alterations, querying them against publicly available databases and generating interactive tools to allow the user to make a swift and efficient analysis of NGS data. In molecular tumor boards, patients are discussed in a limited amount of time. Annotations and tools offered by the *MTPpilot* support clinical decision making. For several years a local version of the *MTPpilot* application has been used at the molecular tumor board of the Comprehensive Cancer Center at the University Hospital Zurich in Switzerland. The software has proven to be useful to analyze in a fast yet comprehensive manner complex NGS panel results, and provide additional information for rare, not trivially interpretable alterations and alterations classified as Variants of Unknown Significance (VUS). By providing the *MTPpilot* as a free web application we want to offer a similar experience to the biomedical community of clinicians. The application is available at the URL www.mtp-pilot.org.

## METHODS

### Database Sources

*MTPpilot* uses at its core a Microsoft SQL database populated with tables from other publicly available databases. Data from the public databases form a reference, evidence and annotation data layer within *MTPpilot*. The web application is implemented with PHP for the backend, and HTML and Javascript for the frontend (Figure 5).

**Figure 5:**
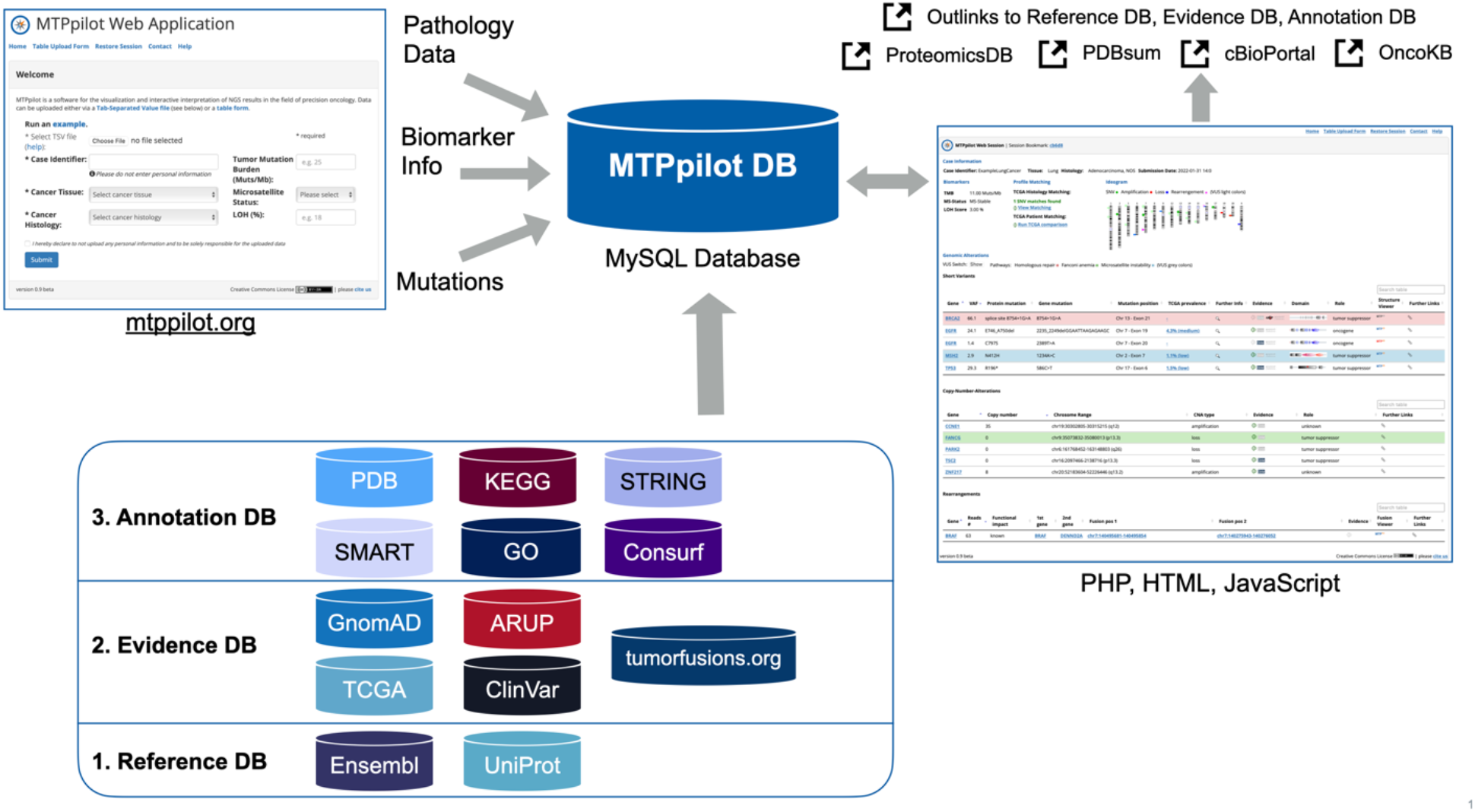
The *MTPpilot* software has at its core a MySQL database with tables holding data from various publicly available databases and a web interface implemented in PHP, HTML and JavaScript. The publicly available databases function either as a reference, evidence or annotation source. Mutation data, biomarker information and pathology data is provided by the user via the *MTPpilot* website and is matched automatically against *MTPpilot* DB. The matching results are presented on a website in a tabular form and extended with interactive visualizations, including and many out links to the reference, evidence, annotation and other databases.

### Reference Databases

Reference databases provide information on the genomic location of genes, transcripts, exons and proteins. For *MTPpilot*, we use the Ensembl database version 105 [21]. Genomic location information for GRCh37 and GRCh38 coordinates including UniProt ID [22] for chromosomes 1 to 22, X, Y, and M were downloaded using the BioMart data mining tool provided on the Ensembl website. Haplotypes and genome patches were ignored.

### Evidence Databases

*MTPpilot* utilizes Evidence databases to assess the pathogenicity of mutations. At the moment, the Evidence databases include TCGA, ClinVar, GnomAD, tumorfusions.org [23] and for BRCA1 and BRCA2 mutations the ARUP database.

For TCGA we downloaded from the TCGA data repository (https://portal.gdc.cancer.gov/repository) MUTECT2 annotated simple nucleotide variation VCF files for 11,037 Whole Exome Sequencing cases covering 33 TCGA projects. In addition, 11,104 Gene Level Copy Number data for the same 33 TCGA projects were downloaded.

From ClinVar, we downloaded 862,195 GRCh37 and GRCh38 variants via the tab-separated file variant_summary.txt.gz that is provided on ClinVar’s FTP server (ftp://ftp.ncbi.nlm.nih.gov/pub/clinvar/tab_delimited/).

GnomAD GRCh37 and GRCh38 variants were downloaded from GATK’s best practice Google Cloud repository (https://storage.cloud.google.com/gatk-best-practices/). GRCh37 variants can be found at <u>/somatic-b37/af-only-gnomad.raw.sites.vcf</u>, while GRCh38 variants are located at <u>/somatic-hg38/af-only-gnomad.hg38.vcf.gz</u>.

As no gene fusion data is available at the TCGA data repository, we took all 20,731 gene fusion events from the Supplementary Table S1 of the recent publication from the tumorfusion.org database (https://doi.org/10.1093/nar/gkx1018).

The ARUP database for BRCA1 and BRCA2 mutations were downloaded directly from the ARUP tables at https://arup.utah.edu/database/BRCA/Variants/BRCA1.php and https://arup.utah.edu/database/BRCA/Variants/BRCA2.php.

### Annotation databases

Annotation databases provide *MTPpilot* with various functional information on mutation. *MTPpilot*’s annotation databases include the Protein Data Bank (PDB), STRING [24] protein interaction database, SMART domain database, GeneOntology (GO) database [25], KEGG pathway database [26] and ConSurf protein conservation database [27].

*MTPpilot* maps mutations on PDB structures to visualize their potential effect on protein functions. The amino acid sequence in PDB structures, however, often deviates from the sequence in protein databases, which is why a simple amino acid number selection in PDB structures is difficult. To circumvent this problem, we pairwise aligned each PDB sequence to its associated UniProt sequence and stored an amino acid mapping table into the *MTPpilot* database.

To highlight the location of a mutation within conserved protein domains, *MTPpilot* maps mutations on the secondary protein structure of the SMART domain database. URLs for retrieving secondary structure images were obtained by the SMART and STRING interaction network developers. The URLs are adjusted at run-time during result page loading to accommodate a red label at the location of the mutation in the protein sequence.

GO information for each gene was retrieved via the Biomart data mining tool as described above. Gene associations to KEGG pathway were retrieved directly from the KEGG Markup Language (KGML) files that are available at the KEGG pathway web sites. The KGML files were downloaded on the 19th March 2019 and processed to extract the pathway name, KEGG pathway ID and gene name. The resulting files were uploaded as an SQL table to the *MTPpilot* DB.

In order to provide information on the evolutionary conservation degree of the mutated amino acid, the ConSurf algorithm was applied on all human canonical protein sequences in UniProt. A copy of the ConSurf software was provided upon request by the ConSurf developers. The software was applied on the UniRef90 database (downloaded on the 29. December 2020) with a maximum number = 300 homologs used ConSurf calculation, 1 iteration of PsiBlast search with an E-score < 0.0001 and the MUSCLE multiple sequence alignment program [28]. Subsequently, the .grades output files of ConSurf were parsed and the conservation scores were saved as a table in the *MTPpilot* DB.

All files generated as described above were further filtered for 1035 relevant genes that are part of Foundation Medicine’s FoundationOneCDx panel [29], Illumina’s TruSight Oncology 500 panel [30], and Thermofisher’s Oncomine Focus and Comprehensive Assay panels [31].

### Implementation of Interactive Visualizations

*MTPpilot* provides various interactive visualisation tools to interact with the mutational data. The most important tools are a chromosome ideogram, TCGA histology matcher, TCGA patient matcher, TCGA prevalence viewer, SMART domain mutation viewer, *MTP3D PDB* viewer and the *MTPfusion* viewer.

#### Ideogram

For drawing a chromosome ideogram to provide a quick summary view of all mutations and mutated chromosomes, *MTPpilot* uses the ideogram.js library version 1.5.0 developed by Eric Weitz (https://eweitz.github.io/ideogram). In the ideogram, short variances are shown as green circles, amplifications as red squares, copy number losses as blue squares and fusions as pink triangles. Variants of unknown significance are shown in the same colors but opaque.

#### TCGA Histology Matcher

For each histology of the TCGA SNV dataset the top 20 mutated genes were calculated, and the cancer histologies categorized into 37 cancer tissues. For each tissue, an additional general histology was generated which contains the data of all other histologies of the tissue (for example the histology "Lung cancer (NOS)” for the tissue "Lung”). The user can first select a tissue, and then one of the associated TCGA histologies that matches best to the uploaded data. The SNV provided by the user are then matched against the top 20 mutated genes from the TCGA dataset. The number of matches are listed in the biomarker and mutational profile section, and the user can open a histogram with the matches highlighted in red via a pop-up.

#### TCGA Patient Matcher

*MTPpilot*’s TCGA patient matcher compares the mutational profile of a tumor board case with *MTPpilot*’s TCGA database. For the comparison, all pathogenic mutations of the tumor board case are matched against all mutations in the TCGA database. A float number score is computed for each match in the format “*n*.*m*”, where the pre-decimal number *n* is the number of identical mutated genes between the tumor board case and a TCGA case and the decimal number *m* is the number of identical mutations. The value of *m* is always equal or smaller to the value *n*, depending on whether all or some mutations are identical between the tumor board case and a TCGA case. Two cases with the same number of identical mutated genes *n*, but different number of identical mutations are ranked such that the case with a larger *m* is higher than the case with the smaller *m*. The score can be found by hovering over the values in the Similarity column. To put the score into context and report the score in percent similarity, we divide each score by the score of the tumor board case. As a result, the tumor board case is always 100% similar to itself, and equal or less similar to the other TCGA cases.

#### TCGA Prevalence Viewer

The prevalence viewer eases the recognition of hotspot mutations in cancer genes by providing a bar plot of mutational frequencies for all amino acid positions ordered from left to right from higher to lower frequencies. The frequencies are computed based on over 1,754,000 SNV data from TCGA. Hot spot regions are typically more frequently mutated than other regions of the gene. To determine a hot spot region, we first excluded all amino acid positions with a single mutation in TCGA. Next, using the frequencies of the remaining positions, we computed the mean and standard deviation frequency for the gene and defined hotspot regions, if their frequency is higher than 10 and higher than the sum of the mean and standard deviation.

#### MTP3D PDB Viewer

To visualize the mutation on protein structures we have implemented the *MTP3D PDB* viewer based on the javascript NGL Viewer library version 2.0.0-dev.39 [32]. The *MTP3D* viewer shows by default the PDB structure in cartoon representation with an opaque visualisation of the molecular surface. Small molecules and HETATM groups are represented as multicolored spheres. Buttons are available to toggle between the structural representations of the protein including cartoon, sticks+balls, spheres and surface representations. Checkboxes allow to hide or show HETATMs, water+ions, all mutations in TCGA or a protein structure with a wild-type amino acid at the mutation side. The latter will show any conformational change inflicted by the mutation.

*MTP3D* will by default show protein structures, which were crystallized with the mutation of the case. If not available, a structure with coordinates of the mutation site, e.g. of the wild type sequence, will be displayed. In any case, a drop-down menu at the top edge of the *MTP3D* viewer allows the user to switch to any other protein structure of the mutated protein. To ease the selection, the PDB title is given next to the PDB Id. A small information panel at the top right corner of the *MTP3D* viewer presents information on the availability of the mutation in the protein structure, the resolution of the PDB structure, the number of residues in the structure, and the percentage of the original protein sequence covered by the PDB structure.

#### MTPfusion Viewer

The *MTPfusion* viewer highlights the location of fusion breakpoints on gene structures. For rendering gene structures, *MTPpilot* uses the Snap.svg javascript library version 0.5.1 (http://snapsvg.io). Exon and intron elements including UTR regions are drawn based on exon, intron and UTR start and end positions stored in *MTPpilot*’s reference Ensembl database (see above). PFAM domains are indicated with additional green boxes labeled with PFAM domain names, whose position is determined by amino acid start and end positions retrieved from the Ensembl database. Whether a fusion event is in-frame or out-of-frame is determined via codon-phase information from Ensembl’s Biomart data mining tool (see above).

## Data Availability

MTPpilot database was build using publicly available data from the Ensembl database version 105, PDB (as retrieved on the 11-Jan-2021), gnomAD (af-only-gnomad.raw.sites.vcf from GATK best practise Google cloud repository), TCGA (as retrieved on 17-Jul-2021), ClinVar (retrieved on 10-Jan-2022), the ARUP Database (as retrieved on 04-Jan-2022), KEGG database (retrieved on 19-Mar-2019), tumorfusions.org (as retrieved on 13-Aug-2021) and ConSurf (computed with UniProt data retrieved on 29-Dec-2020).

https://www.mtppilot.org

## AUTHOR CONTRIBUTIONS

MZ, AK and FMA conceived the software. AK retrieved and processed the data from the publicly available databases and built the MTPpilot database. AK programmed the backend with support of FMA. FMA programmed the frontend with support of AK. JH programmed the TCGA prevalence tool. CP and MW annotated cancer types and cancer histologies for the TCGA histology matching tool. CB and HM provided input for the software content and required functionalities for the tumor board. MZ, AK and FMA wrote the manuscript.

## ACKNOWLEDGEMENTS

We thank Domingo Aguilera for his continuous support in this project.

